# Understanding the Utility of Endocardial Electrocardiographic Imaging in Epi-Endocardial Mapping of 3D Reentrant Circuits

**DOI:** 10.1101/2024.03.13.24304259

**Authors:** Maryam Toloubidokhti, Omar A Gharbia, Adityo Parkosa, Natalia Trayanova, Alexios Hadjis, Roderick Tung, Saman Nazarian, John L. Sapp, Linwei Wang

## Abstract

**Background:** Studies of VT mechanisms are largely based on a 2D portrait of reentrant circuits on one surface of the heart. This oversimplifies the 3D circuit that involves the depth of the myocardium. Simultaneous epicardial and endocardial (epi-endo) mapping was shown to facilitate a 3D delineation of VT circuits, which is however difficult via invasive mapping.

**Objective:** This study investigates the capability of noninvasive epicardial-endocardial electrocardiographic imaging (ECGI) to elucidate the 3D construct of VT circuits, emphasizing the differentiation of epicardial, endocardial, and intramural circuits and to determine the proximity of mid-wall exits to the epicardial or endocardial surfaces.

**Methods:** 120-lead ECGs of VT in combination with subject-specific heart-torso geometry are used to compute unipolar electrograms (CEGM) on ventricular epicardium and endocardia. Activation isochrones are constructed, and the percentage of activation within VT cycle length is calculated on each surface. This classifies VT circuits into 2D (surface only), uniform transmural, nonuniform transmural, and mid-myocardial (focal on surfaces). Furthermore, the endocardial breakthrough time was accurately measured using Laplacian eigenmaps, and by correlating the delay time of the epi-endo breakthroughs, the relative distance of a mid-wall exit to the epicardium or the endocardium surfaces was identified.

**Results:** We analyzed 23 simulated and in-vivo VT circuits on post-infarction porcine hearts. In simulated circuits, ECGI classified 21% as 2D and 78% as 3D: 82.6% of these were correctly classified. The relative timing between epicardial and endocardial breakthroughs was correctly captured across all cases. In in-vivo circuits, ECGI classified 25% as 2D and 75% as 3D: in all cases, circuit exits and entrances were consistent with potential critical isthmus delineated from combined LGE-MRI and catheter mapping data.

**Conclusions:** ECGI epi-endo mapping has the potential for fast delineation of 3D VT circuits, which may augment detailed catheter mapping for VT ablation.

## Introduction

Most life-threatening ventricular tachycardia (VT) episodes involve reentrant circuits facilitated by narrow strands of surviving tissue in areas of patchy scar or at scar borders (1,2). Current analyses of the morphology of reentrant circuits primarily rely on catheter mapping on one surface of the ventricles (epicardium or endocardium). This provides a two-dimensional (2D) simplified view and interpretation of the morphology of the reentrant circuit, assuming all of its critical components to be on one surface.

The spatiotemporal construct of a reentrant circuit, however, is naturally 3D involving mid-myocardium wall. As illustrated in Fig. 1, the electrical current may exit from the protected central isthmus anywhere in the thickness of the myocardial wall. After traveling across the wall, it then breakthroughs to both surfaces of the wall. While observing the earliest site of activation on the surface where measurement is taken (*e.g.*, via catheter mapping), it is important to note that – depending on the 3D construct of the reentrant circuit – the observed surface breakthrough sites may or may not be near the actual exit sites both in transmural depth and surface distances. In an inspiring recent study by Tung et al (3), simultaneous epicardial and endocardial (epi-endo) catheter mapping was used to show that the information on the two surfaces can be combined to facilitate *qualitative* inferences about mid-myocardial activation of 3D reentrant circuits (3). Such knowledge can be important for informing ablation strategies, but unfortunately is mostly missing in the current clinical practice due to the difficulty to obtain simultaneous high-resolution epi-endo catheter mapping, especially for VT episodes where up to 90% of the circuits are too short-lived to permit detailed mapping (2,4).

**Figure 1:**
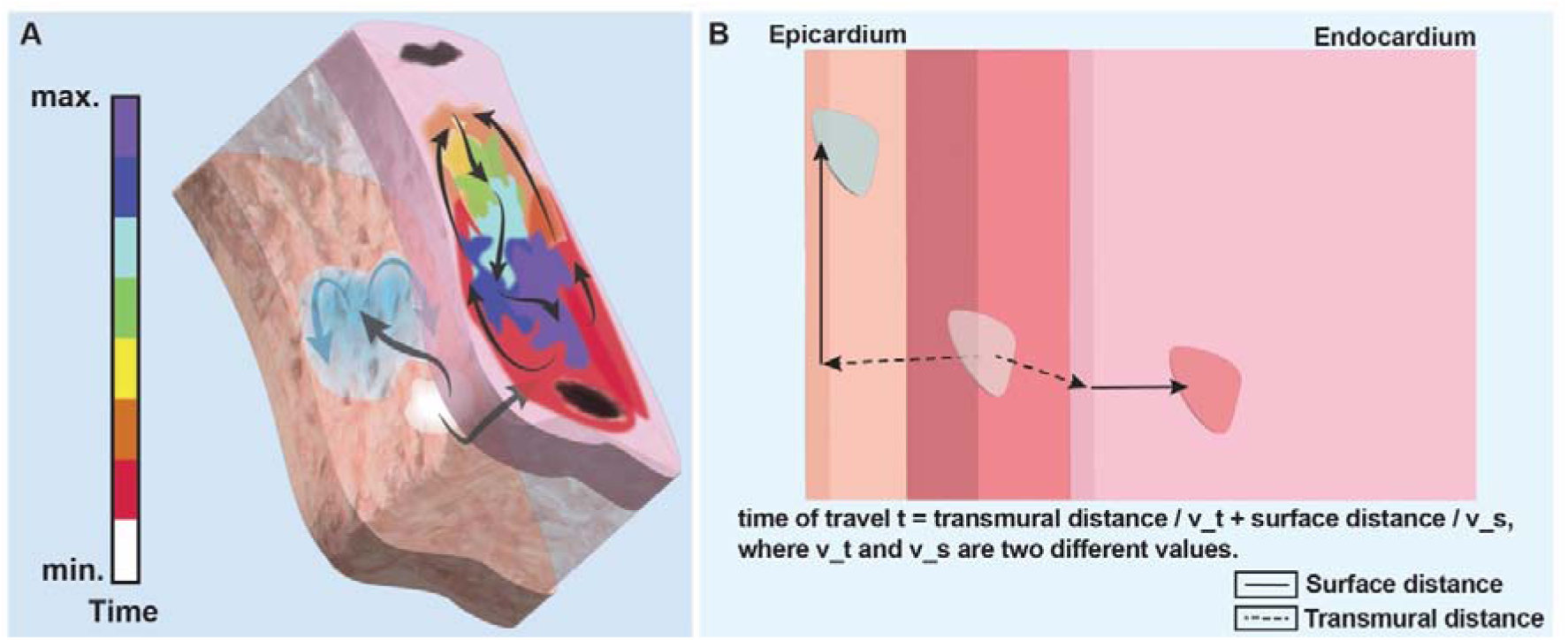
Illustration of a 3D reentrant circuit and its epicardial and endocardial observations. A: A mid-wall exit exhibits as a partial rotation at one surface and a focal activation at the other surface. B. Illustration of a simplified relation between the breakthrough times on both surfaces and the exit site.

Noninvasive electrocardiographic imaging (ECGI) – a family of computational approaches to reconstructing the temporal course of cardiac electrical sources from body-surface electrocardiograms (ECGs) and patient-specific heart-torso geometry – offers a natural candidate to fill this critical gap. However, two critical questions remain. First, despite its widely-explored use for a variety of cardiac arrhythmias such as atrial fibrillation (5) and premature ventricular contraction (6), ECGI techniques have been mainly accepted for their epicardial imaging abilities: the clinical feasibility and utility of endocardial ECGI solutions remain less established despite numerous technical studies (14,15). Second, the study of ECGI in ventricular reentrant circuits have been mostly limited to epicardial solutions including both torso-tank (7,8) and human studies (9–12): notably, a recent study has shown that epicardial ECGI is able to localize the origin of VT circuits with sufficient accuracy to support targeted mapping for ablation (13). Only a small number of studies have investigated the potential of simultaneous epi-endocardial ECGI (14,15) in mapping reentrant VT, although remaining at qualitative inspections of the activation pattern or localization of the site of earliest activation on one of the ventricular surfaces. It has not been yet investigated whether or how epicardial-endocardial ECGI solutions may be combined to elucidate the 3D construct of VT reentrant circuits.

In this study, we will provide answers to these two open questions by examining the feasibility of inferring the 3D construct of a reentrant circuit from what ECGI can reconstruct on both the epicardial and endocardial surfaces of the wall after breakthroughs. The contribution of this study is two-fold. First, with a deep dive into ECGI-reconstructed unipolar electrograms at the endocardium, we show that – while the spatiotemporal amplitude of the reconstructed signals shows significant discrepancies from the ground truth, the phase of the signal in the VT cycle is accurately preserved. We further show that by leveraging Laplacian eigenmaps to quantify the progression of endocardial activation in a two-dimensional space, we were able to Identify the timing of endocardial breakthrough from endocardial ECGI solutions. These findings present one of the first steps towards discovering clinically relevant information hidden within endocardial ECGI solutions, going beyond signal amplitudes. Second, we take a step further built on the results presented in (3) to perform a mechanistic study to use ECGI to infer the *unobserved* mid-wall components of a 3D circuit from the *observed* activity after breakthroughs at the two surfaces of the wall. Following (3), we first show that the gross epicardial and endocardial activation patterns, based on the percentage of circuit activation observed on each surface, can be combined to categorize the reentrant circuit into 2D, 3D, and mid-myocardial constructs.

Due to the fundamental challenges to obtain either clinical (or even experiemntal data) for the transmural morphology of a reentrant circuit, we carry out our investigations in two types of data with different levels of reference data available. First, we leverage 23 previously-published high-fidelity simulations of 3D reentrant circuits virtually induced on post-infarction porcine model (16), to support a mechanistic study where detailed 3D morphology of the reentrant circuits is available. We then extend the investigations to four *in-vivo* animal models where – although 3D mapping data of the reentrant circuit is not available – potential critical isthmus for the reentrant circuits and its transmural depth are estimated from combined analysis of contrast-enhanced magnetic resonance imaging (CE-MRI) and electroanatomic mapping (EAM) data.

Experimental results from the *in-silico* mechanistic study showed that activations observed on the two surfaces – especially the timing and distance of the breakthrough sites – were well correlated with the 3D locations of the exit sites of the reentrant circuits. On these data, combining epicardial and endocardial ECGI activation patterns were able to correctly characterize the categories of 3D reentrant circuits in 83% of the cases, and combining ECGI epi-endocardial breakthrough timing was able to successful differentiate sub-endocardial versus sub-epicardial exit sites in 100% of the cases. In *in-vivo* animal models, the epicardial and endocardial breakthrough time as discovered by ECGI was also to correctly differentiate the one sub-epicardial, one sub-endocardial, and two mid-wall circuits as suggested by the combined MRI-EAM analysis, with the locations of these breakthroughs qualitatively corroborated by MRI and EAM data.

These results provided evidence that ECGI may be a viable tool for providing noninvasive information at the epicardial and endocardial surfaces – especially in the form of the timing and location of the breakthrough sites at these surfaces – that can be combined to infer the 3D construct of a reentrant circuit. This may have a potentil to inform ablation strategies, such as epicardial versus endocardial access or the appropriate penetration depth necessary for an ablation lesion, prior to the procedural.

## Methods

### Epi-Endo Electrocardiographic Imaging

We consider ECGI for reconstructing the time sequence of extracellular potential (*i.e*., unipolar electrograms) throughout the epicardial and endocardial surfaces (15). By representing the bi-ventricular model with a closed surface as illustrated in Fig. 2, the relationship between the heart-surface electrogram and body-surface ECG is governed by a Laplace’s equation defined by the quasi-static electromagnetism (17). When solved numerically on discrete surface bi-ventricular and torso meshes, we obtain a forward matrix ***H*** using open-source SCIRun toolkit (19) that relates unipolar potential at the ventricular surface to body-surface potential though at any time instant. Given a time sequence of, *ϕ_b_*(*t*), we solve, *ϕ_ν_*(*t*) c independently at each time instant by solving the second-order Tikhonov regularization (18)

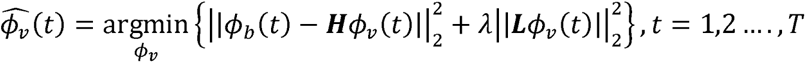

where the first term minimizes the error of fitting body-surface ECG data, and the second term regularizes the solution. Here, the regularization matrix **L** is chosen to be the surface Laplacian operator and the regularization parameter. *λ* is tuned empirically. The inverse calculation was done using a custom Matlab routine.

**Figure 2:**
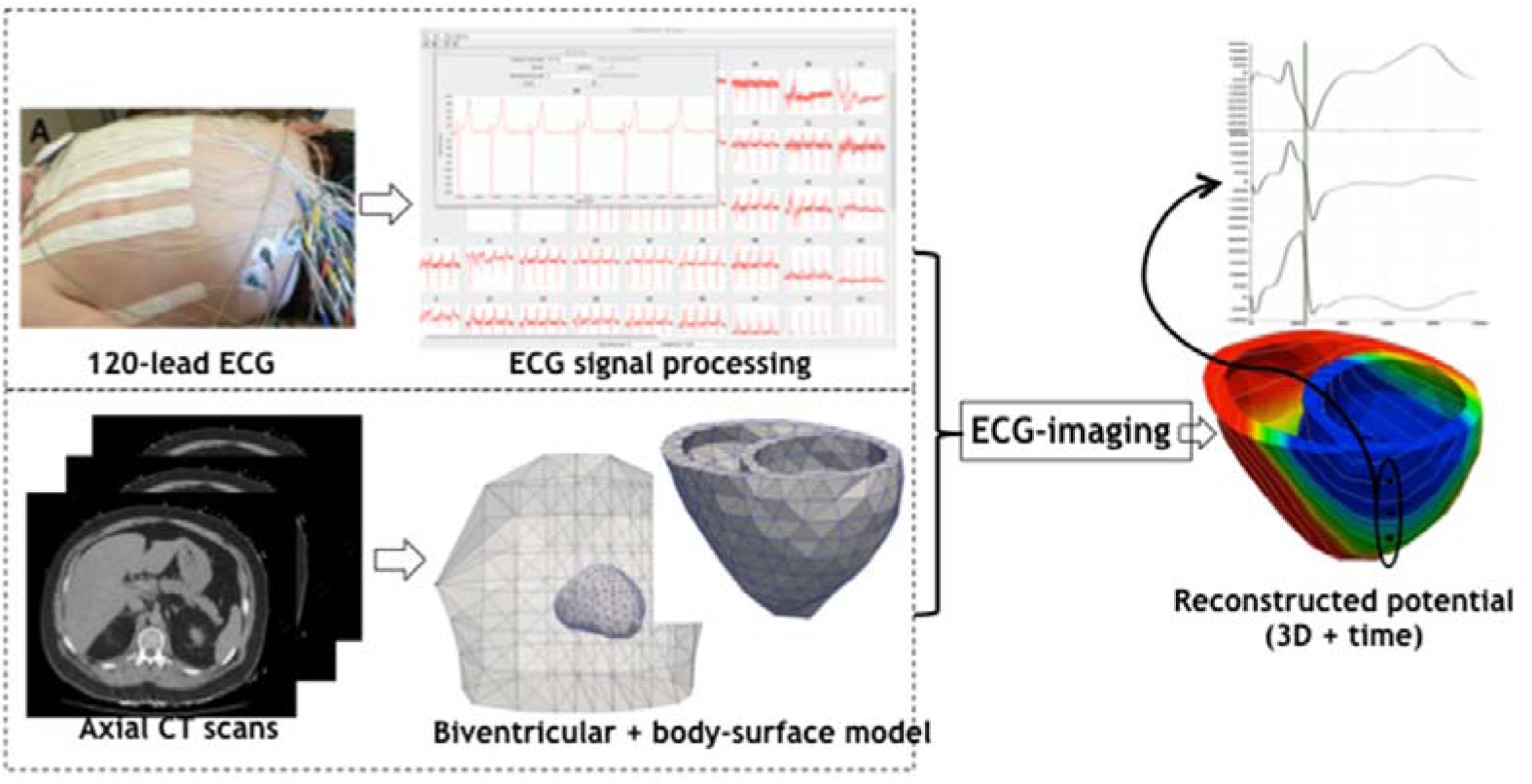
ECGI workflow. Reconstruction of heart electrical potentials from body surface potentials by obtaining body surface recordings from 120 leads placed on the subject’s body and generating heart and torso meshes from MRI images.

### 3D Categorization of Reentrant Circuits

Following (3), we simultaneously quantify the activation isochrones on both the epicardial and endocardial surfaces, and use their relation to categorize the observed reentrant circuit. To extract activation isochronal maps, we first apply the Hilbert transform to heart-surface EGMs to obtain the instantaneous phase signal at each spatial location as described in (20). The activation wave-front at any time instant is then determined as the location with phase = *π*/2 as suggested by (15,21). From the isochronal maps, we locate the region where rotations of activation can be seen and identify the portion of rotation – out of the full cycle of VT – that is visible on each surface. The activation gap, as defined in (3), is the period of time when activation of the rotation is missing from a surface. If a rotation is not at least partially visible on a surface, the activation is considered focal on that. Finally, we combine the isochronal maps on both surfaces to categorize the 3D characteristics of a reentrant circuits following the definition used in (3):

- 2D circuits: A complete rotation without any activation gap (all isochronal colors) is seen on one surface, and focal activation is seen on the other surface.
- 3D non-uniform circuits: Partial rotations with activation gaps are observed on one or both surfaces.
- 3D uniform circuits: Complete rotations are observed on both surfaces. This was not observed in the data used in this study.
- Mid-myocardial circuit: Focal activation is observed on both surfaces.

### Identifying the Timing and Location of Endocardial Breakthrough Sites

Epicardial ECGI solutions are generally consistent in reconstructing the amplitude of the potential signals in the time domain, as illustrated in Fig. 3A. In comparison, endocardial ECGI solutions tend to create artificial macroscopic rotation patterns that exhibit large discrepancy from the actual potential signal, suggesting significant errors in the reconstruction of the signals in the time domain as illustrated in Fig. 3B. Does this suggest that ECGI solutions are completely invalid on the endocardial surfaces?

**Figure 3:**
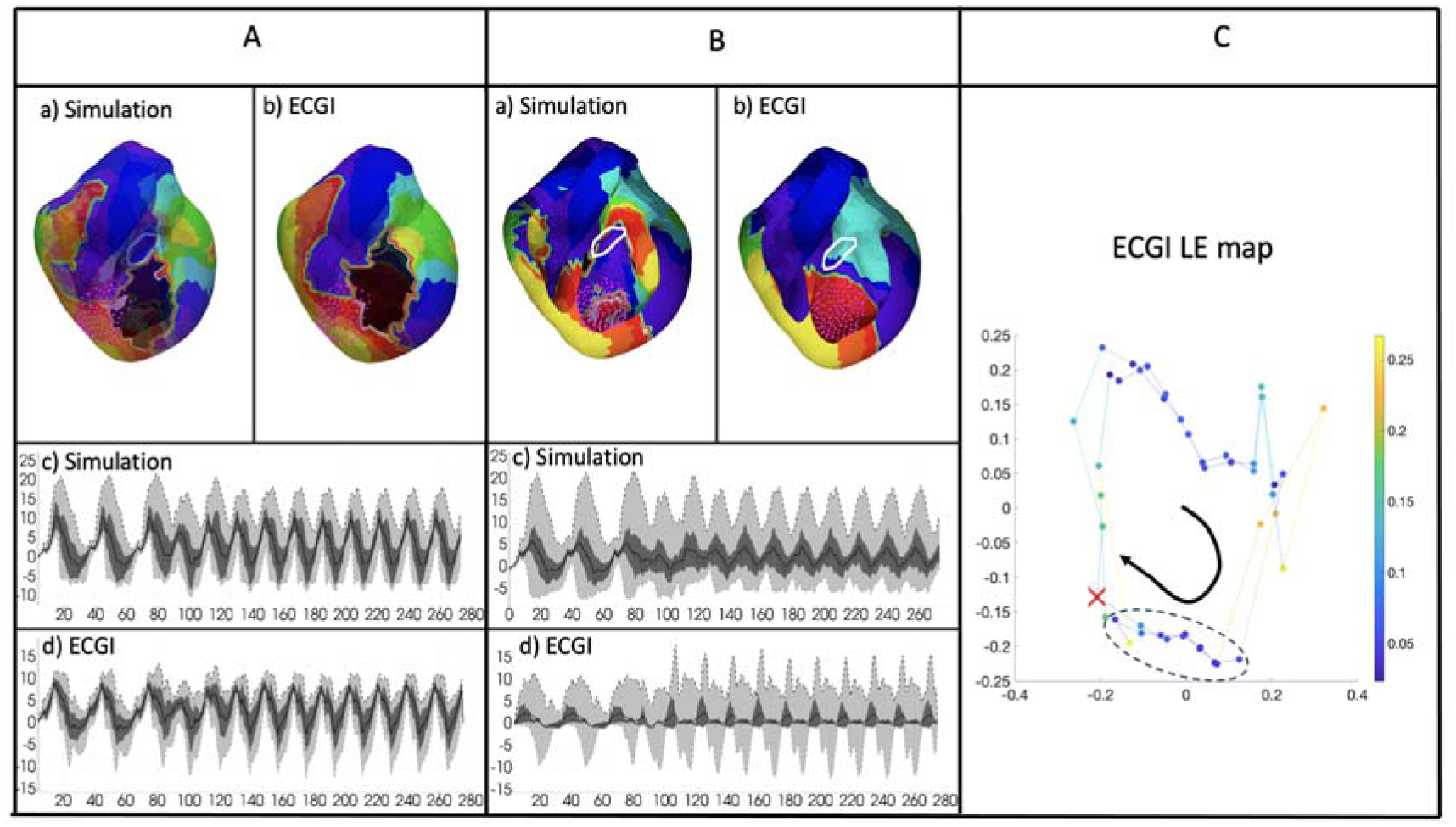
Analyzing the ECGI solutions on epicardium and endocardium. A) Comparing ECGI pattern and amplitude on epicardium B) Comparing ECGI solution pattern and amplitude on endocardium C) example of identifying the endocardial breakthrough time from the Laplacian eigenmap of the ECGI solution.

A closer look at the endocardial ECGI solutions reveals that, while the amplitude of the signal *ϕ_ν_*(*t*) are incorrectly reconstructed at each time instant t, a notable pattern of slow progression followed by fast progression is consistently observed in each VT cycle in the reconstructed *ϕ_ν_*(*t*) sequence on the endocardium. We hypothesize that the phase of this high dimensional signal *ϕ_ν_*(*t*) – its progression in the periodic cycle of the VT beats – is captured in ECGI solutions, and that the time instant following the slow progression can be identified as the time of breakthrough on the endocardium.

To extract the phase of *ϕ_ν_*(*t*) for discreate snapshots of the temporal signal at t = 0, 1, …T, we leverage Laplacian eigenmaps (LE) (26) as a nonlinear dimensionality reduction method to obtain a low-dimensional representation of the spatial-temporal signal sequence of ECGI solutions on the endocardium. The primary objective of using LE is to explore and uncover potential patterns in the spatiotemporal activation sequences over time by visualizing this activity in a lower dimensional space. The method constructs a graph using pairwise distances between data points and a heatmap kernel, to capture relationships between the values of the neighbor nodes. Subsequently, the graph Laplacian matrix is computed and subjected to Singular Value Decomposition (SVD). Through the SVD, the significance of the manifold’s coordinates is determined and ranked, from which we then use directions associated with the two largest eigenvalues to preserve features of the original signals in a significantly reduced space.

The resulting two-dimensional coordinate space, termed the “LE space”, represents a manifold or a trajectory of the endocardial ECGI solution pattern over time where, as illustrated in Fig. 3C, the reconstructed full endocardial potential at time frame t is represented by a unique position on the manifold in the two-dimensional LE space and each “loop” represents the progression of *ϕ_ν_*(*t*) within a monomorphic VT cycle. As shown, the duration of slow progression can be characterized with narrowly spaced points whereas the duration of rapid progression can be characterized with sparsely spaced points. We thus use the distance between the 2D points, i.e., *ν*(*t*) = ||*d*(*k*) − *d*(*k* − 1)||^2^1)||, as a surrogate measure of the speed of progression (color coded in Fig 3C), and identify the time of endocardial breakthrough as the time instant at which *ϕ_ν_*(*t*) exits from the duration of slow progression, as marked by the red cross in Fig 3C.

### From Surface Breakthrough Sites to Exit Sites Beneath the Surface

We further investigate if the breakthrough sites observed on the two surfaces bounding the myocardial wall can be combined to reveal the location of circuit exits inside the wall. Fig. 1B illustrates our intuition: that the relative timing and location of the breakthrough sites on the two surfaces of the wall reflects how the electrical current travels to the two surfaces after exiting from the scar. To quantify this, we proceed in two steps.

As a primary focus, we attempt to correlate the delay time of the two breakthroughs to the relative proximity of the mid-wall exits to either of the two surfaces bounding the wall. We primarily focus on differentiating whether a mid-wall exit is closer to the epicardium or the endocardium, where the latter can be either left endocardium or right endocardium depending on the location of the reentrant circuits. To do so, we record the time difference between the epicardial and endocardial breakthroughs dt = *t_epi_* – *t_endo_*, and study the use of dt for differentiating sub-epicardial/sub-endocardial exit sites. The reference sub-epicardial versus sub-endocardial location of the actual mid-wall exit is determined based on the actual mid-wall location of the exit in simulation data, and by the MRI and/or ablation evidence in animal and human data. We hypothesize that a negative dt, with an earlier breakthrough at the endocardium, indicates a sub-endocardial exit site, a positive dt with an earlier epicardial breakthrough indicates a sub-epicardial exit site, and a dt close to 0 indicate a mid-wall exit site. We confirm this hypothesis by the epicardial-endocardial breakthrough time identified in the simulated reentrant circuits, and then test the ability of epi-endocardial ECGI solutions in capturing this.

## Materials and Data

### Simulation Reentrant Circuits

The use of simulation data, while associated with limitations pertaining to validity in comparison to real data, provides unique availability of detailed 3D data of the reentrant circuits that is not possible in current experimental or clinical settings. Here, we utilize reentrant circuits virtually induced on detailed ventricular model with high-resolution scar morphology from a previous study (16). As detailed in (16), simulation data were generated on eight chronically infarcted porcine hearts. Detailed models of the intact large animal ventricles were constructed from *in-vivo* MRI images, with image-based fiber orientation and detailed scar geometry obtained at a voxel size of 0.03125 mm^3^. Electrical wave propagation was modeled by the monodomain formulation, and the simulations were performed using the software package CARP (CardioSolv, LLC) on a parallel computing platform (16). Monomorphic VTs were induced in all the hearts using a clinical S1-S2-S3 programed electrical stimulation protocol (22), applied from 27 pacing sites selected on the basis of a modified American Heart Association segment designation. A total of 23 sustained VTs (lasting for at least 2s after the last pacing stimulus) were induced and used in this study. For each sustained VT, the pathway of the reentrant circuit in the form of a string loop was identified by connecting seed points along the fastest part of the 3D activation of the circuit as described in (16),

To simulate body-surface ECGs corresponding to each reentrant circuit, each animal heart was placed in a human torso model consisting of a triangular mesh with 120 vertices representing the positions of surface ECG leads. Epi-endocardial extracellular potential was simulated from each reentrant circuit solving the Poisson’s equation underlying the quasi-static electromagnetism using a custom software (23). This served as the cardiac source model and used to generate the 120-lead body-surface ECG using the forward operator specific to each heart-torso pair constructed using the SCIRun software (19). The surface ECG was added with 20-dB Gaussian noise and input for ECGI reconstruction of epi-endo EGMs.

### Chronic Post-Infarction Animal Models of Reentrant Circuits

Four swine models, weighed 35-45 kg, were used in a study protocol in accordance with the Johns Hopkins University Institutional Animal Care and Use Committee (16,24). Prior to the invasive procedure of MI creation, swine were anesthetized with mechanical ventilation using a combination of tiletamine, zolazepam, ketamine, and xylazine and maintained under sedation using 1-2 % isoflurane. In all animals, MI was created by inserting a guiding catheter into the left coronary artery and occluding the left anterior descending (LAD) coronary artery using an angioplasty balloon for two hours.

At approximately eight to ten weeks post-MI, DCE-CMR was performed in all swine models using a 3-T scanner (Prisma, Siemens Healthcare) in 20-30 minutes after injection of Gadopentetate dimeglumine (0.20 mmol/kg, Magnevist, Bayer, Leverkusen, Germany). A free-breathing navigator-gated three-dimensional inversion recovery T1w sequence was used with typical imaging parameters as: inversion time = 400 ms, flip angle = 25°, repetition time = 5.4 ms, echo time = 2.7 ms, reconstructed pixel size = 1.1 x 1.1 x 1.1 mm with interpolation in the slice direction, 12 segments per imaging window, GRAPPA acceleration factor (R) = 2, FOV = 300 x 220 mm, matrix = 272 x 200, bandwidth = 200 Hz/Pixel, scan time = 15-20 min.

Within one week after DCE-CMR, the electrophysiological study was performed using the NavX mapping system (EnSite Velocity, St. Jude Medical). Prior to the electrophysiology study, 18 strips of 120 disposable radiolucent electrodes were placed on the swine torso following the standard Dalhousie mapping protocol (25). Approximately ten surface electrodes were removed to accommodate the placement of NavX pads. The rest of the electrodes remained attached throughout the electrophysiology study to record surface ECG during induced VT.

In heparinized animals, endocardial mapping of the left ventricle was first performed during sinus rhythm via a retrograde approach using a duodecapolar (20-electrode) catheter (interelectrode spacing = 1-2.5-1 mm, electrode size = 1 mm, AfocusII, St. Jude Medical, Minnetonka, MN) with the NavX mapping system (EnSite Velocity, St. Jude Medical). In addition, a decapolar catheter was advanced from the right jugular vein to the coronary sinus. Programmed ventricular stimulation was performed to induce VT from two right ventricular sites (outflow tract and apex), with up to three extrastimuli decremented to ventricular refractoriness or 250 ms at two drive trains (600 and 400 ms). If VT is induced, mapping during VT was attempted.

## Results

### Results on Simulated 3D Reentrant Circuits

The following information were extracted from each reentrant circuit, both simulated “ground truth” and epi-endocardial ECGI solutions: 1) the 3D categorization of reentrant circuits based on qualitative isochrone analysis, 2) the time of epicardial and endocardial breakthrough, and 2) the location of epicardial and endocardial breakthrough (when possible). On the simulated “ground truth” of the reentrant circuit, the central isthmus (if observable) and exits were also identified in 3D to differentiate sub-epicardial, sub-endocardial, and mid-wall circuit exits.

Below, we first describe the general behavior of ECGI solutions in reconstructing the VT morphology at the epicardial and endocardial surfaces. We then delve into examining the extent to which quantitative metrics – especially the time and location of the surface breakthrough sites – may be extracted from these surface reconstructions to inform 3D location of the reentrant circuits, both in simulated *ground truth* and in ECGI reconstructions.

#### Epicardial ECGI solutions

Across all 23 cases, on the epicardium, ECGI was able to reconstruct the electrograms with a reasonable accuracy of spatial correlation coefficients of 0.89 ± 0.03, temporal correlation coefficients of 0.90 ± 0.02, and a relative mean squared errors (RMSE) of In all 23 cases, a clear site of the earliest activation could be observed at the epicardium, and was selected as the site of epicardial breakthrough. This resulted in a good accuracy in localizing epicardial breakthrough sites, with 14.17 ± 10.78 mm Euclidean distance error in spatial location and 0.6 ± 0.55ms error in timing for sub-epicardial cases, and 9.39± 5.67mm in spatial location and 0.66 ± 0.59ms in timing for sub-endocardial cases, as summarized in Fig. 4. This suggests that the quality of ECGI epicardial solutions was little effected by the intramural location of a reentrant circuit. Diving deeper into the reconstructed epicardial solutions, we noted several limitations. First, when complete circuit including the protected activity within the critical isthmus can be observed in the simulated epicardial data, ECGI reconstruction typically missed such local protected activation shown Fig. 5A. Second, in a small number of cases, ECGI artificially “closed the loop” of the reentrant circuit when the simulated circuit was only partially observed on the epicardium, as illustrated in Fig. 5B. Finally, even as ECGI closely captured the epicardial breakthrough sites, these sites may be far away from the actual exit if the intramural circuits are further beneath the epicardium, as illustrated in Fig. 5C. As a result of these traits, epicardial ECGI alone – despite good performance in capturing gross activation patterns and localizing the site of breakthrough – may have limited ability to inform the 3D construct of the reentrant circuit and its critical isthmus beneath the surface.

**Figure 4:**
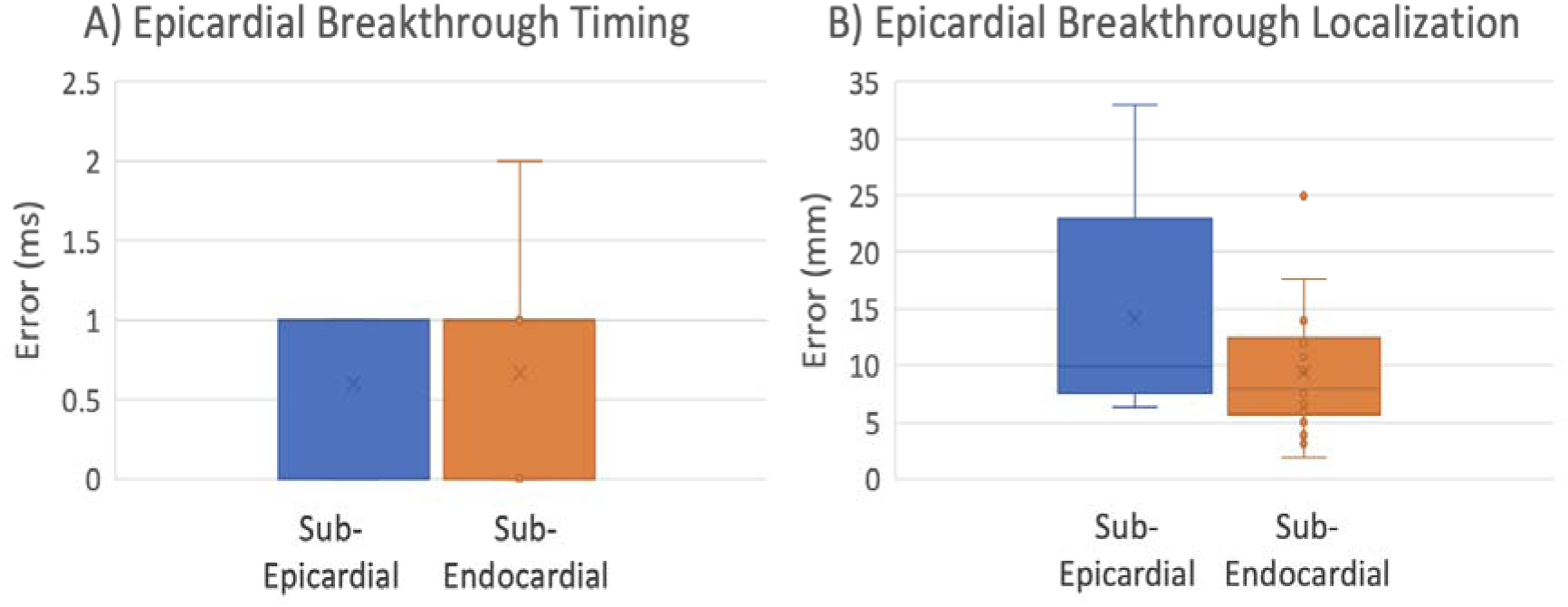
Timing (A) and Localization (B) error of Epicardial-breakthrough categorized by sub-epicardial and sub-endocardial groups.

**Figure 5:**
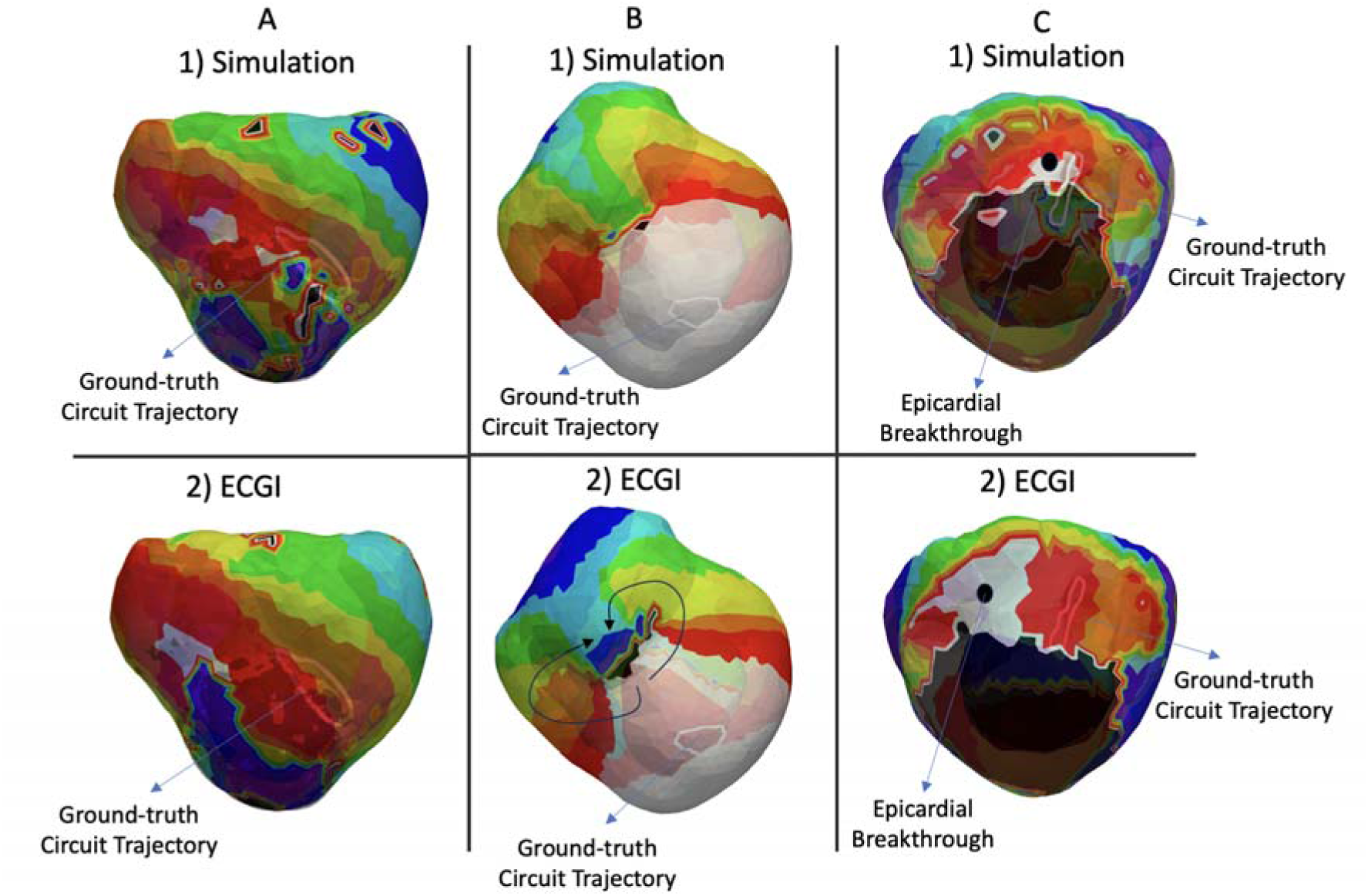
(A) Missing isthmus in ECGI (B) Artificially closing the loop (C) Epicardial breakthrough far from the circuit’s exit site.

#### Endocardial ECGI solutions

Across the 23 cases, ECGI solutions on the endocardium was in general poorer in reconstructing the gross activation patterns compared to its epicardial counterpart, with a spatial correlation coefficient of 0.30 ± 0.11, temporal correlation coefficients of 0.46 ± 0.14, and a relative mean squared errors (RMSE) of. In all cases, there was no evident site of the earliest activation on the endocardium. Instead, an artificial macroscopic rotation pattern was consistently created in all cases, as illustrated in both example A and example B in Fig. 6.

**Figure 6:**
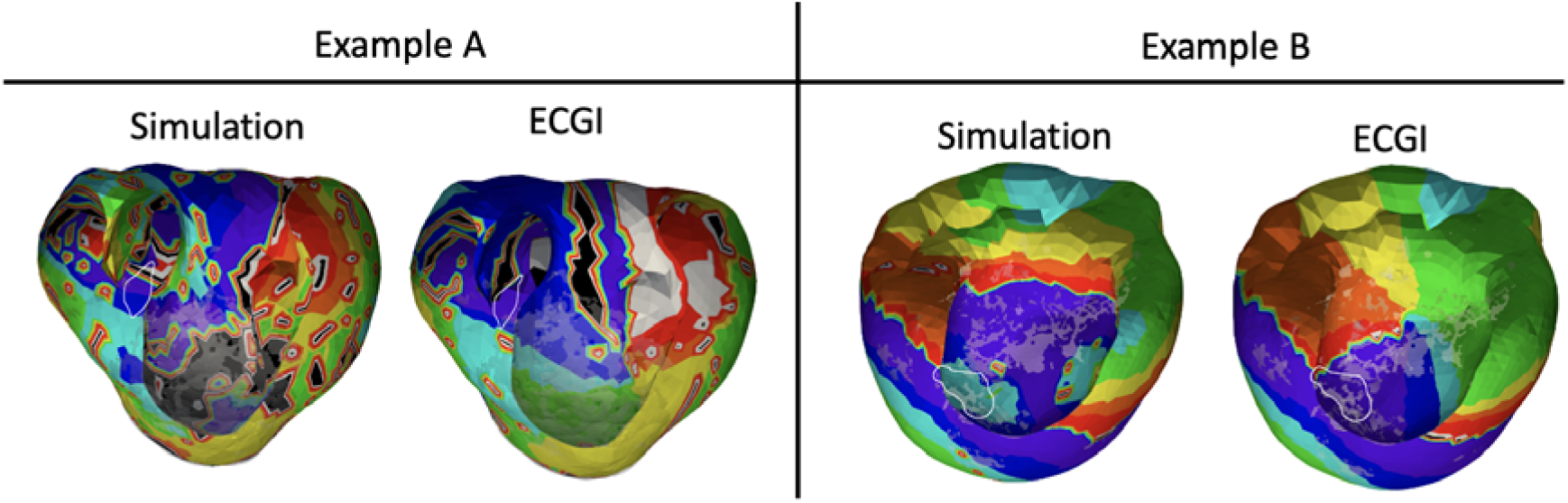
Illustration of the artificial macroscopic rotation pattern created on the endocardial surface by ECGI solutions.

Despite the dismal picture the above observations may suggest for endocardial ECGI solutions, accurate timing for detecting endocardial breakthroughs was obtained with LE-based embedding of the temporal trajectory of endocardial activations as described earlier: over the 23 cases, the error in timing endocardial breakthroughs = 0.66±0.59 for sub-endocardial cases and 0.6 ±0.55 for sub-epicardial cases. In terms of locating the endocardial breakthrough sites, no clear signature was discovered in the majority of the cases. Below, we examine the utility of these endocardial breakthrough measures – despite the overall low fidelity of activation patterns – in informing the 3D construct of reentrant circuits when combined with epicardial measures.

#### 3D Categorization of Reentrant Circuits

Fig. 7A summarizes the 3D categorization of the 23 simulated reentrant circuits. Out of the 23 circuits, 2D circuits were observed in 26% of the cases (n=6) on the endocardium, whereas 3D circuits were found in 74% of the cases (n=17). 23% (n=4) of these 3D circuits were partially observed on only one surface, and one of the circuits was mid-myocardial. None of the reentrant circuits was 3D transmurally uniform. In ECGI reconstruction of these reentrant circuits as summarized in Fig. 7B, 22% were 2D endocardial (n=5), and the rest (78%) were categorized as 3D circuits (n=18). 33% of the 3D circuits were partially observed on only one surface (n=6), and two circuits were reconstructed to be mid-myocardial. None of the ECGI-reconstructed reentrant circuits was 3D transmurally uniform either.

**Figure 7:**
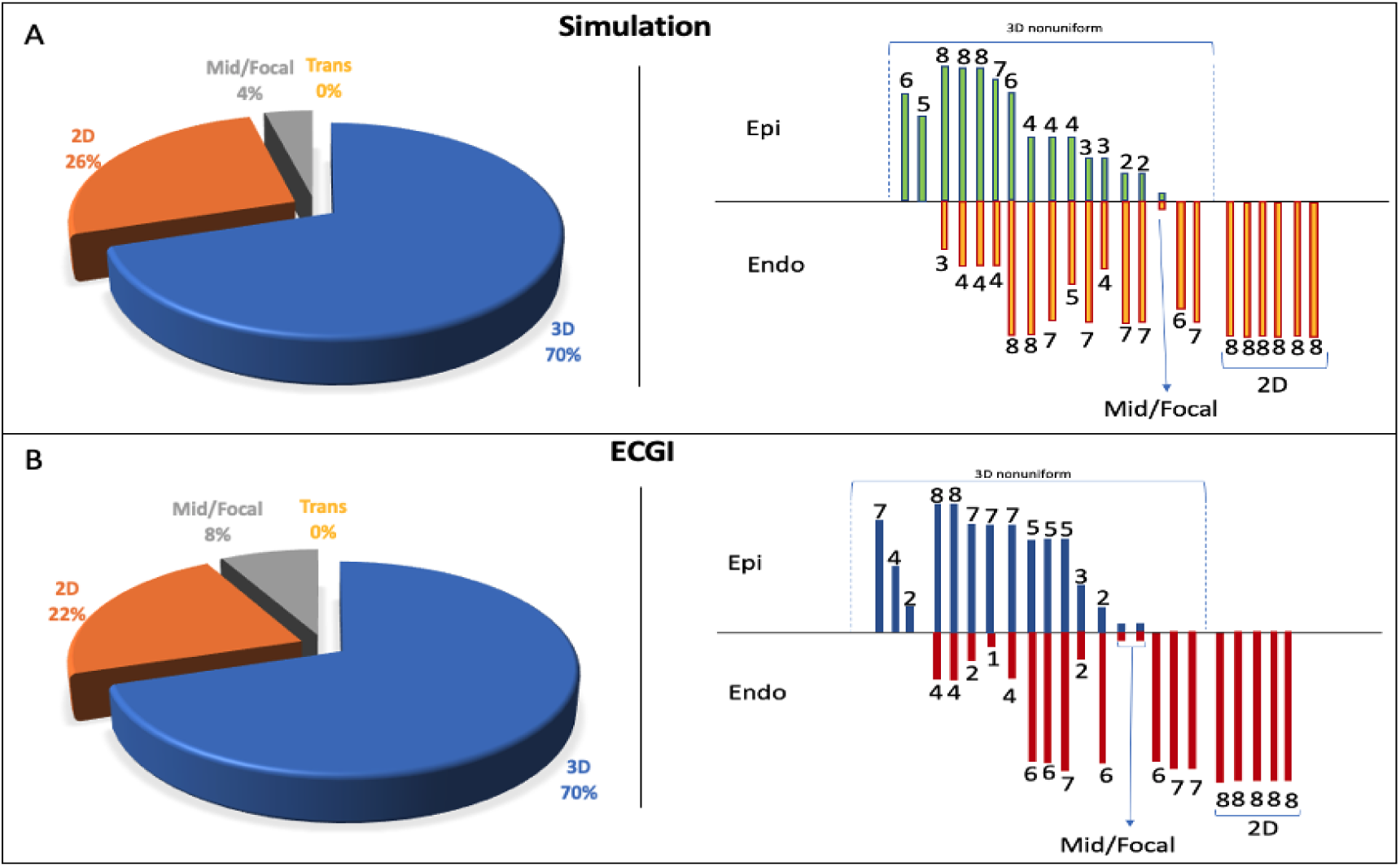
Inferring the 3D category of reentrant circuits using epi-endocardial activation patterns.

Fig. 8A details case-by-case evaluations of ECGI’s ability to recognize the 3D categorization of reentrant circuits against the simulated ground truth. As shown, ECGI reconstruction correctly preserved the 3D categorization of the reentrant circuits in 82% of the cases (n=19). Out of six 2D reentrant circuits, four (66%) were correctly reconstructed by ECGI. Out of the 17 3D reentrant circuits, 15 (88%) were correctly reconstructed by ECGI. In the four incorrectly reconstructed cases, one mid-myocardial circuit was reconstructed to be 2D endocardial, one endocardial circuit was reconstructed to be mid-myocardial and one reconstructed to be 3D nonuniform, and one 3D non-uniform circuit was reconstructed to be mid-myocardial. Fig. 8B-C summarizes the sensitivity,specificity, and confusion matrix of ECGI in reconstructing 2D and 3D non-uniform cases (mid-myocardial and 3D uniform cases were excluded out of the calculation because of the small number or absence ofexamples).

**Figure 8:**
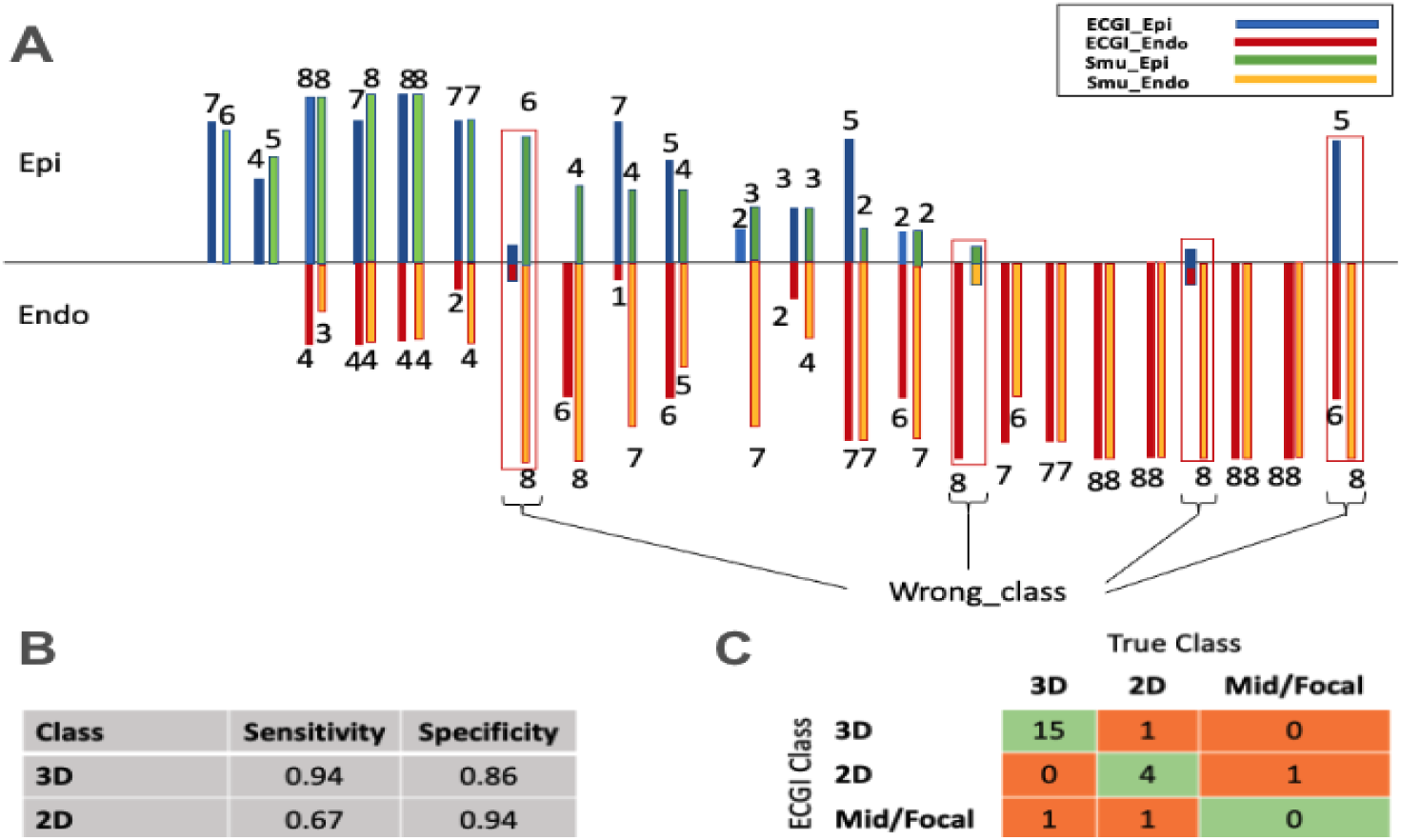
Case-by-case comparison of ECGI recognition of 3D category of reentrant circuits versus simulated ground truth.

#### Differentiating sub-endocardial vs. sub-epicardial exits by the timing of epicardial and endocardial breakthroughs

Fig. 9 summarizes the quantitative results of using the delay between epicardial and endocardial breakthroughs in all circuits to differentiate the seven sub-epicardial and 16 sub-endocardial exits, where the latter could be either left or right endocardium as summarized earlier. In simulated data, all reentrant circuits with subendocardial exits were all associated with an epicardial breakthrough lagging the endocardial breakthrough; circuits with sub-epicardial exits were all associated with an epicardial breakthrough preceding the endocardial breakthrough. This was consistently preserved by ECGI in all cases in terms of the sign of the epi-endocardial breakthrough timing delay, as show in Fig. 9 with an absolute error of 1.65 ± 1.46. This provides initial evidence that the relative order of epi-endocardial breakthroughs in ECGI solutions can consistently be used to differentiate whether the circuit exit was closer to the epicardial or endocardial surfaces.

**Figure 9:**
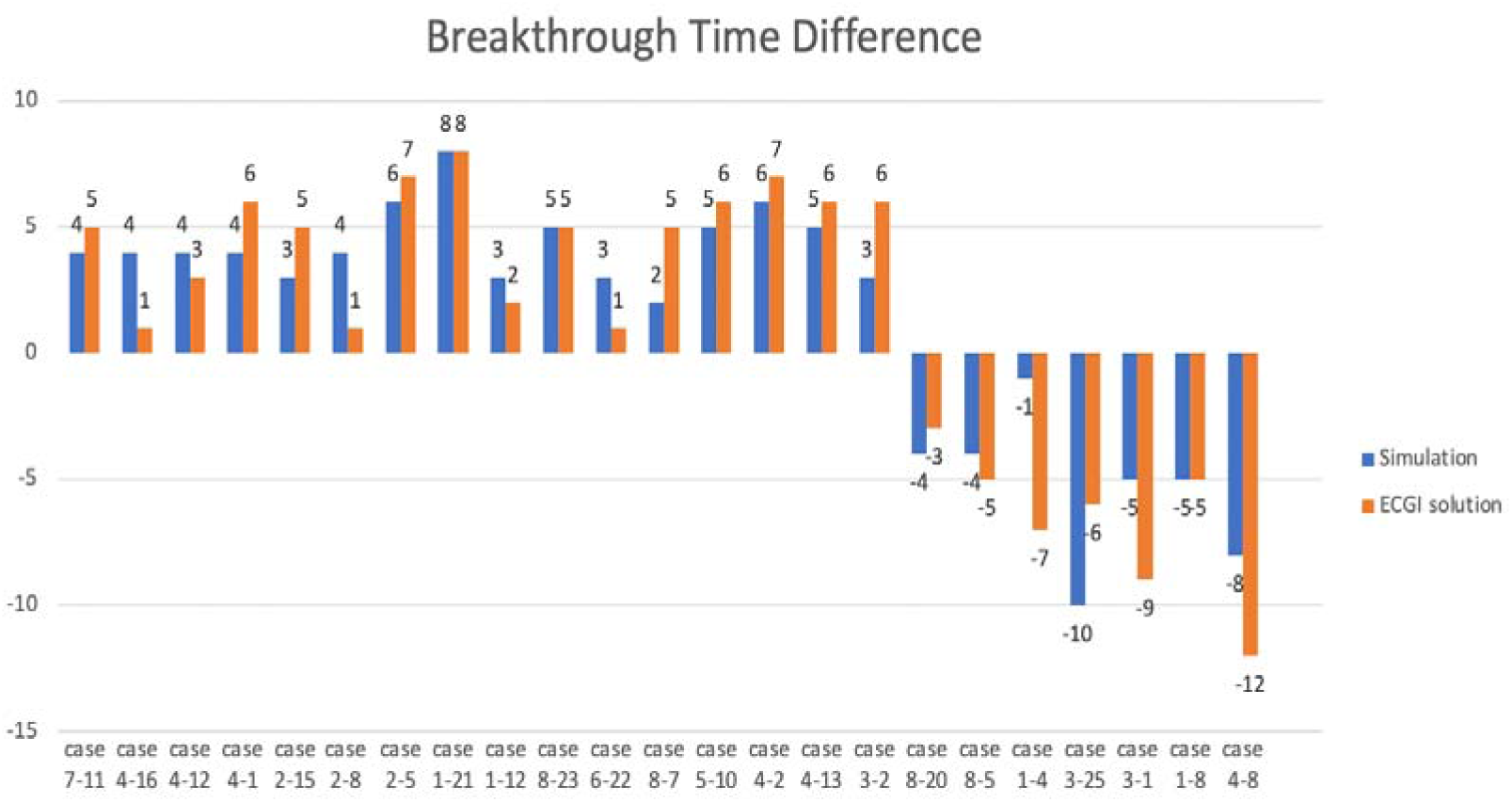
using the delay between epicardial and endocardial breakthroughs to identify the closer surface to the exit site.

### Results on Chronic Post-Infarction Animal Models of Reentrant Circuits

A total of four reentrant VT circuits induced in four animal models were studied. From the DCE-MRI images, ADAS-VT software (ADAS 3D, Galgo Medical, Barcelona, Spain) was used to delineate possible slow-conducting channels within the myocardial scar. In the meantime, *in-vivo* EAMs were analyzed to identify local abnormal EGMs such as double potential, delayed potential, and fractionated potential obtained during sinus rhythm, and mid-diastolic potential or activation maps obtained during induced VT. These two sources of data combined provided reference data of the locations of potential critical isthmus for each of the induced VT circuits. As summarized through Figs 10–13, despite the limited number of cases, a variety of intramural distributions spanning from sub-endocardial, mid-wall, to sub-epicardial isthmus were observed among the four reentrant circuits. The analyses of ECGI results and MRI/EAM data are carried out by independent operators.

**Figure 10:**
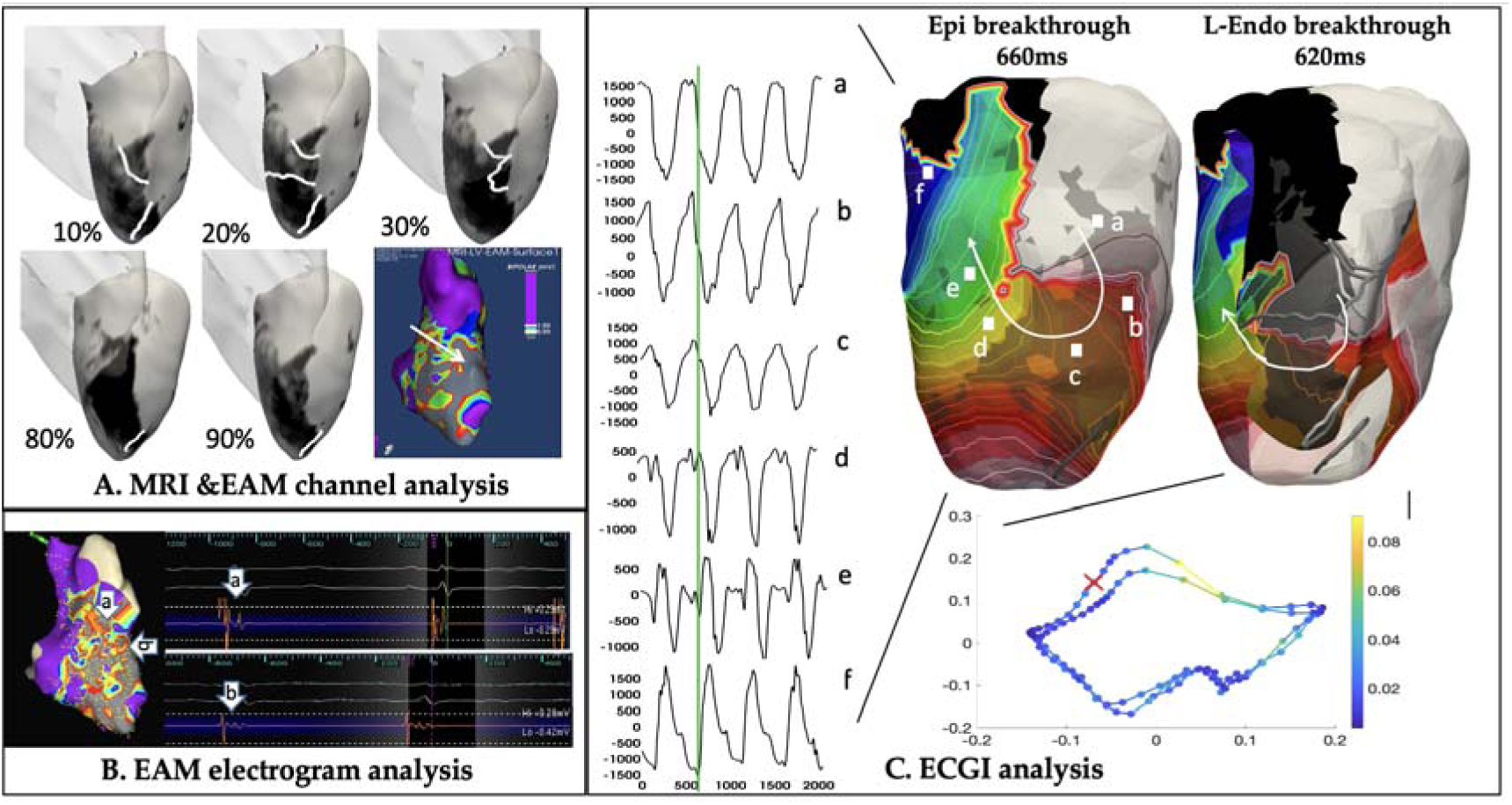
MRI, EAM and ECGI analysis of animal data, case1 with a sub-endocardial anterior septal circuit.

#### Case 1 – sub-endocardial anterior septal circuits

In case 1, DCE-MRI identified conducting channels primarily identified at 10-30% layer of the LV septum at the anterior-septal region, corroborated by the higher-voltage channels detected within the low-voltage area in native-rhythm voltage mapping (Fig 10.A). Fractionated potential was further detected at septal mid-basal-anterior wall during native rhythm electroanatomical mapping (Fig 10.B). In ECGI solutions, the epicardial activation map suggested a breakthrough at mid anterior-septal region of the LV followed by a local clockwise rotation of activation as illustrated by the activation map and selected electrocardiograms (Fig 10.C). ECGI endocardial breakthrough was identified by LE at 40 ms prior to the epicardial breakthrough, followed by also a clockwise rotation with a spatially meandering anchor around the same area on the endocardium (Fig 10.C). This epi-endocardial breakthrough delay suggested a sub-endocardial circuit exit located at mid-basal anterior-septal region of the LV, consistent with the potential channels revealed by combined MRI-EAM analysis.

#### Case 2 – sub-epicardial apical circuits

In case 2, DCE-MRI suggested dense subendocardial scar and potential channels were only detected at 80-90% sub-epicardial layer of the LV (Fig. 11.A). On the EAM, delayed potential was detected around apical septum during native-rhythm mapping, while mid-diastolic and early-systolic potential were detected around apical septum during VT (Fig. 11.B). In ECGI solutions, epicardial activation suggested a breakthrough at the apical region of the LV septum followed by a focal apex-to-base activation as illustrated by the activation map and selected electrocardiograms (Fig. 12C). ECGI endocardial breakthrough was identified by the LE at 40-50 ms after the epicardial breakthrough, with a counterclockwise rotation anchored on the mid-septum of the LV endocardium (Fig. 11C). This epi-endocardial breakthrough delay suggested a sub-epicardial critical channel, located at apical septum of the LV. This was consistent with one of the only two DCE-MRI derived channels at 90% layer (*i.e*., sub-epicardial layer) of the LV septum and the EAM data.

**Figure 11:**
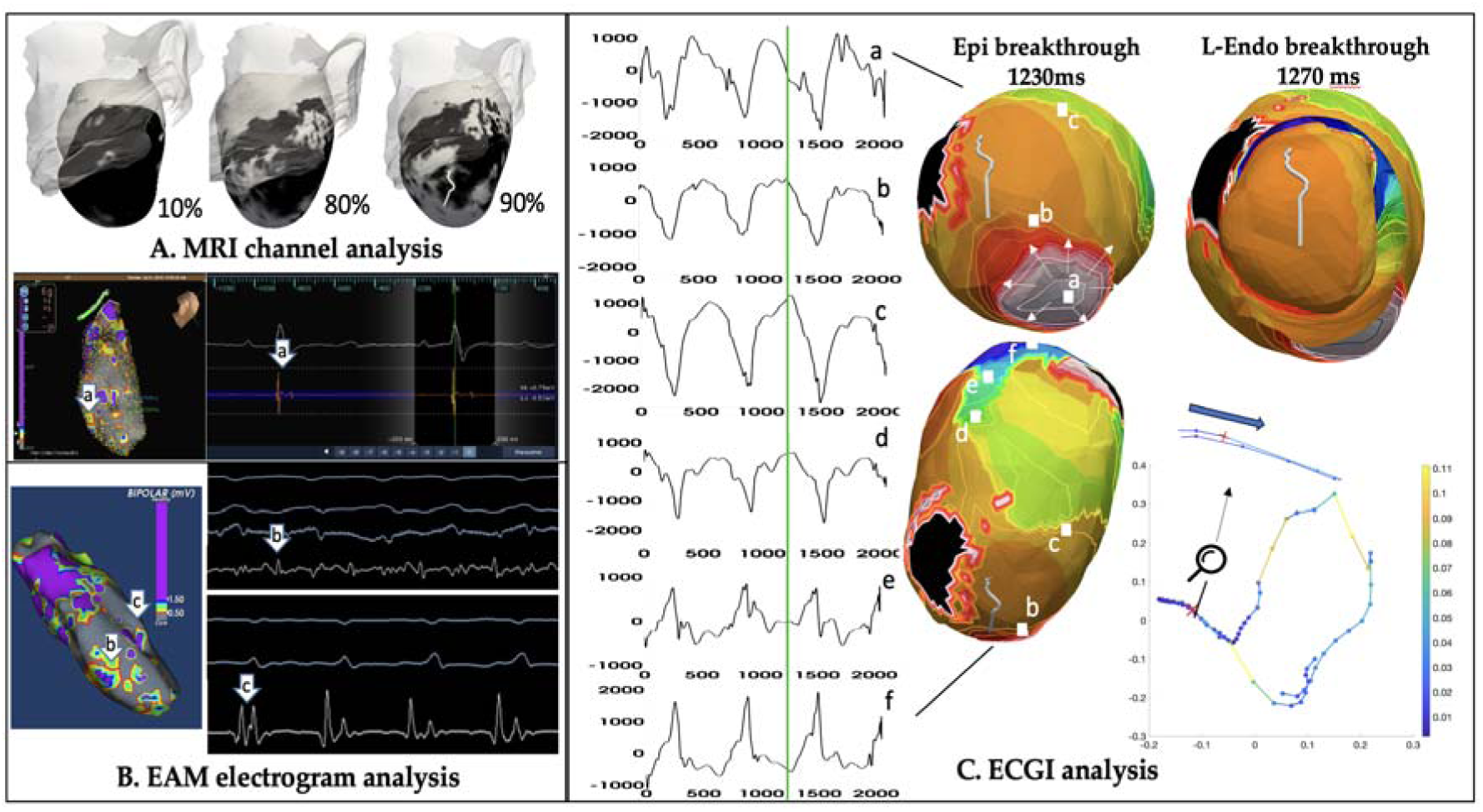
MRI, EAM and ECGI analysis of animal data, case2 with a sub-epicardial apical circuit.

**Figure 12:**
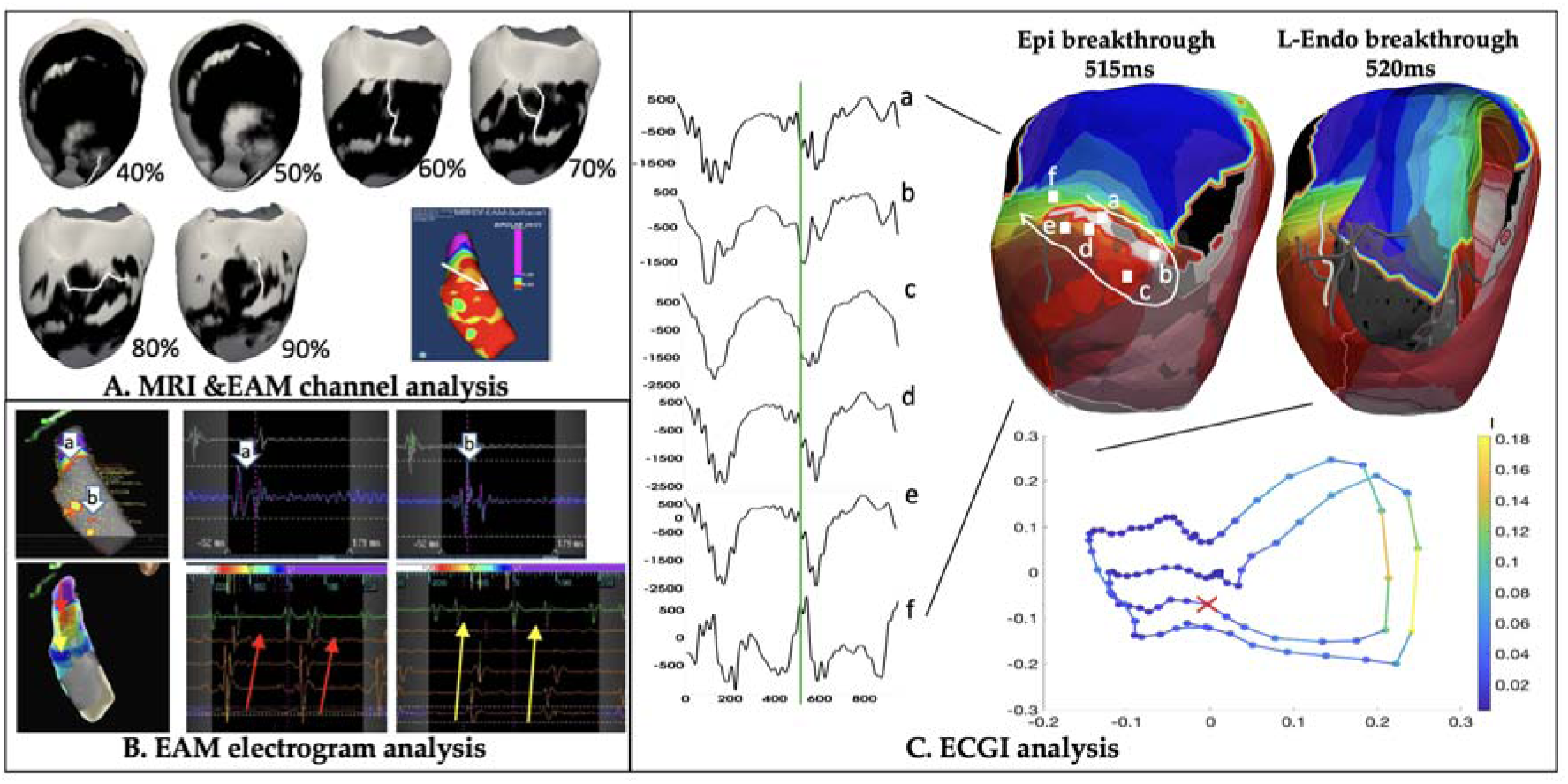
MRI, EAM and ECGI analysis of animal data, case3 with a mid-wall mid-basal anterior-septal circuit.

#### Case 3-4: – Mid-wall mid-septum and anterior circuits

In case 3, DCE-MRI suggested apical channels at the mid-layer (40-50%) of the LV, and mid-basal lateral-anterior region (60-90%) of the LV septum – the latter consistent with higher-potential channel revealed within the low-voltage region during native-rhythm voltage mapping (Fig 12 A). Additionally, double potential was detected at basal and mid-anterior LV during native-rhythm mapping, while mid-diastolic potential around the same region was detected during VT (Fig 12 B). In ECGI solutions, epicardial solutions suggested a breakthrough at mid-basal anterior-septal region of the LV followed by a local clockwise rotation, as illustrated by the activation map and selected electrocardiograms (Fig 12 C). ECGI endocardial breakthrough was identified at 5 ms after epicardial breakthrough, with a counter-clockwise rotation with a meandering anchor in mid-to-apical septal region of the LV endocardium. This epi endocardial breakthrough delay suggested a mid-wall to sub-epicardial circuit exit located at mid-basal anterior-septal region of the LV, consistent with a series of potential channels identified at 60-90% layer channels identified by combined MRI-EAM analysis.

In case 4, DCE-MRI again suggested relative dense scar with basal lateral-anterior channels, corroborated by the relatively higher-voltage channel identified within the low voltage area during native-rhythm voltage mapping (Fig. 13A). VT activation map obtained by EAM suggested an early activation at mid-anterior region of the LV septum and an apex-to-base activation pattern at the inferior region of the LV (Fig. 13B). In ECGI solutions, epicardial activation showed a breakthrough at mid-basal lateral-anterior region of the LV septum, leading to a local clockwise rotation near the mid-basal anterior-septal region as illustrated by the activation map and selected electrocardiograms (Fig. 13C). ECGI endocardial breakthrough was identified by the LE almost simultaneously with the epicardial breakthrough, with a clockwise rotation and a meandering anchor at a similar location to that on the epicardium. This epi-endocardial breakthrough delay suggested a mid-wall critical channel located at basal lateral-anterior region of the LV septum, consistent with the combined MRI-EAM analysis. Furthermore, note that in this case, the endocardial activation map also showed an early activation at mid-anterior region of the LV septum and an apex-to-base rotation on the inferior region, as that mapped during VT *in-vivo*.

**Figure 13:**
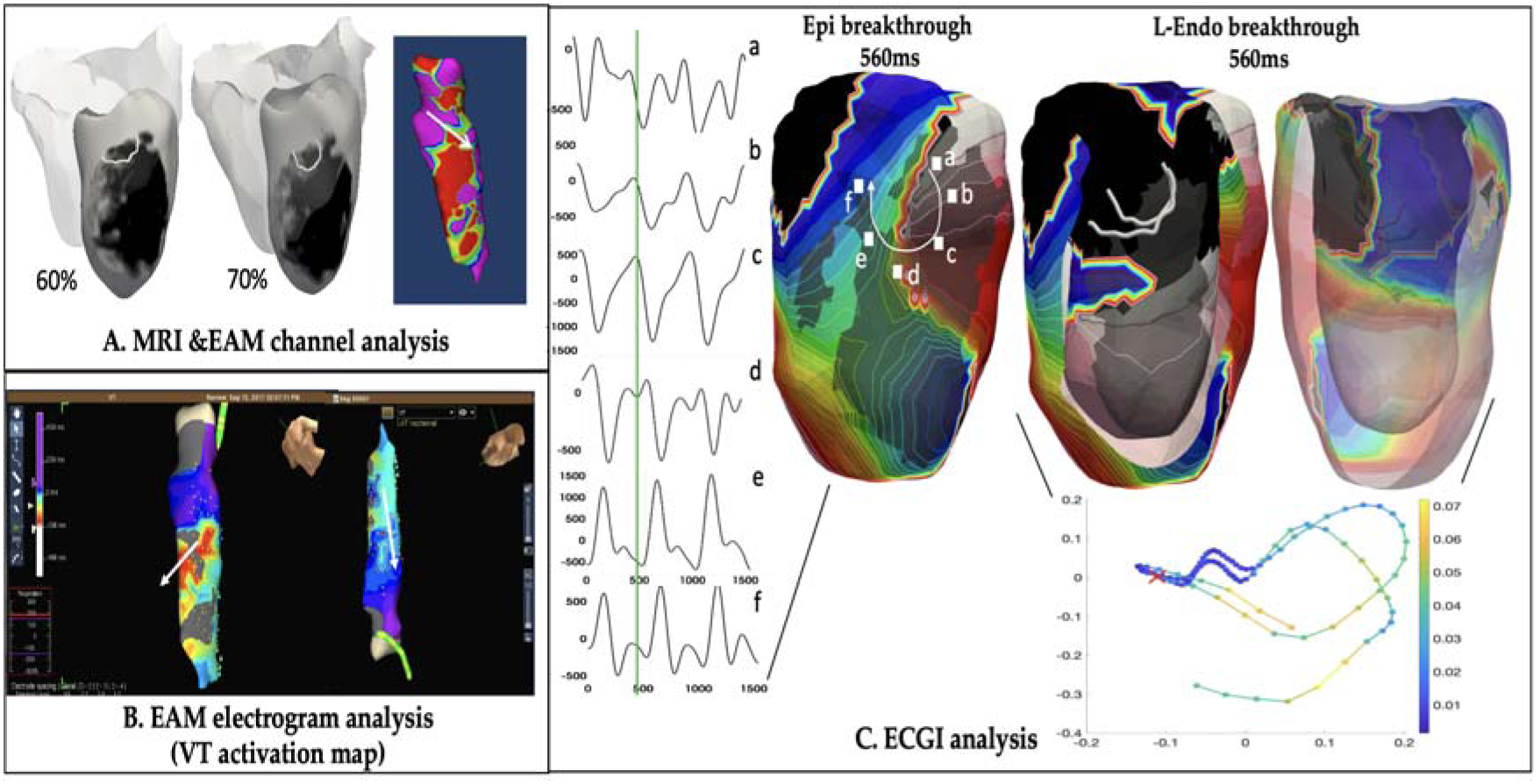
MRI, EAM and ECGI analysis of animal data, case4 with a mid-wall circuit located at basal lateral-anterior region of the LV septum.

Overall, across the four reentrant circuits, it was observed that epicardial ECGI solutions revealed breakthrough sites and general rotation patterns qualitatively consistent with the general location of MRI-EAM suggested sites of critical isthmuses. Details on the activation maps, however, were not sufficient to reveal further information about the potential entrance sites of the local reentrant circuits nor the exact pinpointing of the exit sites, potentially because the signals of the local reentrant circuit was dominated by the global activation pattern throughout the ventricles. Validity on the activation pattern of ECGI endocardial solutions or breakthrough sites was less conclusive. Despite these limitations, the relative timing of epi-endocardial ECGI breakthroughs consistently revealed the proximity of the potential circuit exits to the epicardial or endocardial surfaces, as supported by the combined MRI-EAM analyses – suggesting a potential clinical value for epi-endocardial ECGI in informing ablation strategies.

## Conclusions

While the use of simulation data warrants further translations to experimental or clinical data, it enabled mechanistic investigations of the relation between detailed 3D morphology of reentrant circuits and their observations on epi-endo surfaces. The extension to *in-vivo* animal model and human subject data further provided a proof of concept that simultaneous epi-endo mapping, especially the timing and location of epi-endocardial breakthrough sites, may provide important information for inferring the 3D morphology of the reentrant circuits. ECGI, as a noninvasive technique for rapid and simultaneous epi-endo mapping, may play an important role in revealing the hidden components of a 3D reentrant circuits that is currently not possible. This has potential to enable further 3D mechanistic studies of VT morphologies or guiding ablation strategies in a way that is not possible with current practice.

## Data Availability

Both the simulation and the in-vivo animal data used in this study have been previously used in other published works as referenced in the manuscript and are available upon request.

